# Characteristics and Risk Factors for Hospitalization and Mortality among Persons with COVID-19 in Atlanta Metropolitan Area

**DOI:** 10.1101/2020.12.15.20248214

**Authors:** Nathaniel Chishinga, Neel R. Gandhi, Udodirim N. Onwubiko, Carson Telford, Juliana Prieto, Sasha Smith, Allison T. Chamberlain, Shamimul Khan, Steve Williams, Fazle Khan, Sarita Shah

## Abstract

**Background:** We present data on risk factors for severe outcomes among patients with coronavirus disease 2019 (COVID-19) in the southeast United States (U.S.).

**Objective:** To determine risk factors associated with hospitalization, intensive care unit (ICU) admission, and mortality among patients with confirmed COVID-19.

**Design:** A retrospective cohort study.

**Setting:** Fulton County in Atlanta Metropolitan Area, Georgia, U.S.

**Patients:** Community-based individuals of all ages that tested positive for SARS-CoV-2.

**Measurements:** Demographic characteristics, comorbid conditions, hospitalization, ICU admission, death (all-cause mortality), and severe COVID-19 disease, defined as a composite measure of hospitalization and death.

**Results:** Between March 2 and May 31, 2020, we included 4322 individuals with various COVID-19 outcomes. In a multivariable logistic regression random-effects model, patients in age groups ≥45 years compared to those <25 years were associated with severe COVID-19. Males compared to females (adjusted odds ratio [aOR] 1.4, 95% confidence interval [CI]: 1.1-1.6), non-Hispanic blacks (aOR 1.9, 95%CI: 1.5-2.4) and Hispanics (aOR 1.7, 95%CI: 1.2-2.5) compared to non-Hispanic whites were associated with increased odds of severe COVID-19. Those with chronic renal disease (aOR 3.6, 95%CI: 2.2-5.8), neurologic disease (aOR 2.8, 95%CI: 1.8-4.3), diabetes (aOR 2.0, 95%CI: 1.5-2.7), chronic lung disease (aOR 1.7, 95%CI: 1.2-2.3), and “other chronic diseases” (aOR 1.8, 95%CI: 1.3-2.6) compared to those without these conditions were associated with increased odds of having severe COVID-19.

**Conclusions:** Multiple risk factors for hospitalization, ICU admission, and death were observed in this cohort from an urban setting in the southeast U.S. Improved screening and early, intensive treatment for persons with identified risk factors is urgently needed to reduce COVID-19 related morbidity and mortality.

## INTRODUCTION

Since SARS-CoV-2, the novel coronavirus that causes coronavirus disease 2019 (COVID-19), was detected in the United States (U.S.) in January 2020, it has spread rapidly across the nation with 2.8 million cases and 130,000 deaths as of July 5, 2020.(1, 2) As testing increases and transmission continues, it is critical to understand the epidemiology of this disease to inform clinical practice and prevention strategies. Early data from the US has suggested more severe disease among older persons and those with comorbidities.(3) The most common comorbidities among US COVID-19 patients include hypertension, diabetes, obesity, renal disease, lung disease, and immunosuppression.(4-6) In addition, persons who are non-Hispanic black represent 23% of COVID-19 related deaths where race is known,(7) despite comprising 13.4% of the U.S population.(8) Although hospitalization rates have been reported as higher among black patients, results have been mixed when evaluating black race as a risk factor for death after adjusting for covariates.(9)^-^(10) To date, there have been limited community-based data from the southeastern U.S. where a larger proportion of the population is black, and where there is a disproportionate burden of chronic health conditions and poor access to care.(11)

By August 6, 2020, the state of Georgia had a cumulative estimate of 204895 COVID-19 cases of which 19230 (9.4%) were in Fulton County, with most of these cases residing in the city of Atlanta.(12) Recent estimates show that Fulton County, with an estimated population of over 1 million persons, has a high proportion of non-Hispanic blacks (four times higher than the national average), 13.5% of whom are living in poverty.(8) This epidemiologic context is similar to other jurisdictions in the southeastern U.S.; however, there have been limited inferential data from population-based cohorts on risk factors associated with COVID-19 morbidity and mortality from this region. Therefore, understanding the clinical disease spectrum and risk factors for severe disease can inform public health programs and clinical providers in this highly affected geographic region. Here we report on risk factors associated with hospitalization, ICU admission, and death among COVID-19 patients from Fulton County, Georgia.

## METHODS

### Design, Setting and Participants

This study was a retrospective cohort study of individuals diagnosed with laboratory-confirmed SARS-CoV-2 infection from March 2 to May 31, 2020, who resided in Fulton County (situated in Atlanta Metropolitan Area), Georgia. A laboratory confirmation for SARS-CoV-2 was defined as a positive result of real-time reverse transcriptase–polymerase chain reaction or antigen test.

### Data Source

Data were extracted from the State Electronic Notifiable Disease Surveillance System (SENDSS). SENDSS is an electronic database developed by the Georgia Department of Public Health that is used to track patients with notifiable diseases, including case investigation of COVID-19 patients across the state of Georgia. The extracted data for each case included in this study were: date of first SARS-CoV-2 positive specimen collection, age, gender, race and ethnicity, medical comorbidities, residence in a long-term care facility (LTFC), hospitalization, intensive care unit (ICU) admission, and death. “Other chronic diseases” was a variable in SENDSS used to encompass other comorbidities not itemized. Where applicable, dates related to hospitalization, discharge and death, as well as length of stay in the ICU, and type of respiratory support in the ICU (invasive mechanical ventilation, extracorporeal membrane oxygenation) were also extracted and used to determine hospitalization or ICU admission status for records with missing data.

In order to have complete case investigations and reporting to the surveillance system, we censored the cohort follow-up time to May 31, 2020. We allowed for a 20-day lag to June 20, 2020, for completion of case investigations in SENDSS before extracting the data for analysis.

### Statistical Analysis

Baseline characteristics were presented as medians with interquartile ranges (IQR) for continuous variables and as frequencies with proportions (%) for categorical variables. In standard regression analyses, the assumption is that the individual observations are independent of each other. However, this assumption is not always tenable, especially in congregate settings like LTCFs where infections are correlated. Clusters of cases from LTCFs have been shown to contribute to high proportions of hospitalizations and COVID-19-related deaths.(13) Therefore, in order to account for correlations within LTCFs, we used random effects models.

We fitted logistic regression random effects models for each variable and outcome, with LTCFs as random effects. Variables from the unadjusted random effects models that were statistically significant, i.e., null value of the odds ratio (OR=1.0) not contained within the 95% confidence interval (95% CI), and race/ethnicity as *a priori*, were included in the multivariable random effects models. In the multivariable random effects models we obtained adjusted odds ratios (aOR) and 95% CI for factors associated with hospitalization, ICU admission, death (all-cause mortality), and severe COVID-19 – a composite of hospitalization or death.

We further fitted a multivariable logistic regression random effects model that included race/ethnicity and gender as an interaction term. We used the likelihood ratio test to test if there was evidence of interaction between these two variables in the severe COVID-19 random effects model.

To account for missing covariate and outcome data, we analyzed missing data for each covariate as an unknown category. We further performed a sensitivity analysis where we replaced all participants with “unknown” information on comorbidities with “no” to bias away from the observed effect and then fitted a multivariable logistic regression random effects model for risk factors associated with severe COVID-19. In order to determine if missingness for outcome data was random, we also fitted multivariable logistic regression random effects models that compared (i) age (as a continuous variable) among those with missing outcome data with those without missing outcome data, and (ii) gender among those with missing outcome data with those without missing outcome data.

A two-sided p-value of <0.05 or null value not contained within the 95% CI were considered as statistically significant. Statistical analyses were performed in Stata software version 15.1 (Stata Corp, College Station, Texas, U.S.), unless otherwise stated.

### Ethical Considerations

This activity was determined to be consistent with public health surveillance activity as per title 45 code of Federal Regulations 46.102(l)(2). It was approved by the Emory University institutional review board with a waiver of informed consent.

## RESULTS

Between March 2 and May 31, 2020 there were 2820 individuals diagnosed with SARS-CoV-2 infection with hospitalization-related information (yes, no) available (median age 54 years, IQR 37-69; range 0-104 years; 52.0% female); 1650 patients with ICU-related information available (median age 51 years, IQR 36-65; range 0-103 years; 50.8% female), and 1969 patients with mortality-related information available (median age 56 years, IQR 38-70; range 0-104 years; 50.6% female). There were 2,851 patients with either hospitalization or mortality-related information available, of whom 949 (33.3%) had severe COVID-19.

We found no statistical difference in age or gender between those with missing information and those with available hospitalization-related information (OR for age 1.0, 95%CI 1.0-1.0; OR for gender 1.0, 95%CI: 0.9-1.2); available ICU-related information (OR for age 1.0, 95%CI 1.0-1.0; OR for gender 1.0, 95%CI: 0.9-1.2); available mortality-related information (OR for age 1.0, 95%CI 1.0-1.0; OR for gender 1.2, 95%CI: 1.0-1.3). We therefore excluded cases that had missing outcome information in our analyses and assumed that they were missing at random.

### Characteristics of COVID-19 Cases

Among the 2820 patients with available hospitalization-related information, 888 (31.5%) were hospitalized; of those who were hospitalized, 440 (49.5%) were ≥65 years old, 473 (53.3%) were male, and 632 (71.2%) were non-Hispanic black (**Table 1**). Of the 1650 patients with available ICU-related information, 239 (14.5%) were admitted to ICU; of these, 131 (54.8%) were ≥65 years old, 150 (62.8%) were male, and 180 (75.3%) were non-Hispanic black. Of the 1969 patients with mortality-related information available, 295 (15.0%) had died as of May 31, 2020; of those who died, 239 (81.0%) were ≥65 years old, 163 (55.3%) were male, and 219 (74.2%) were non-Hispanic black (**Figure 1, Table 1**).

**Table 1.**
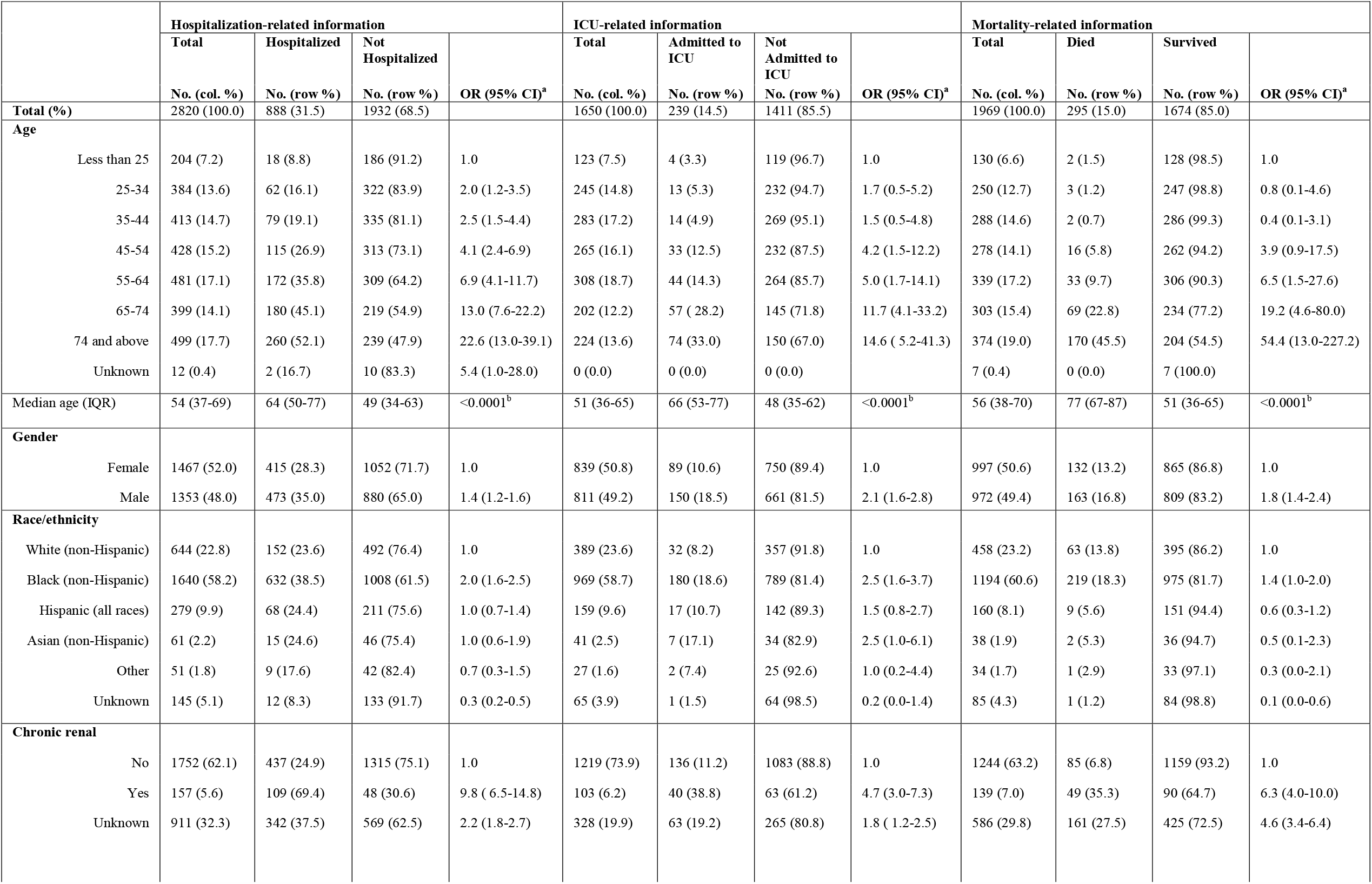
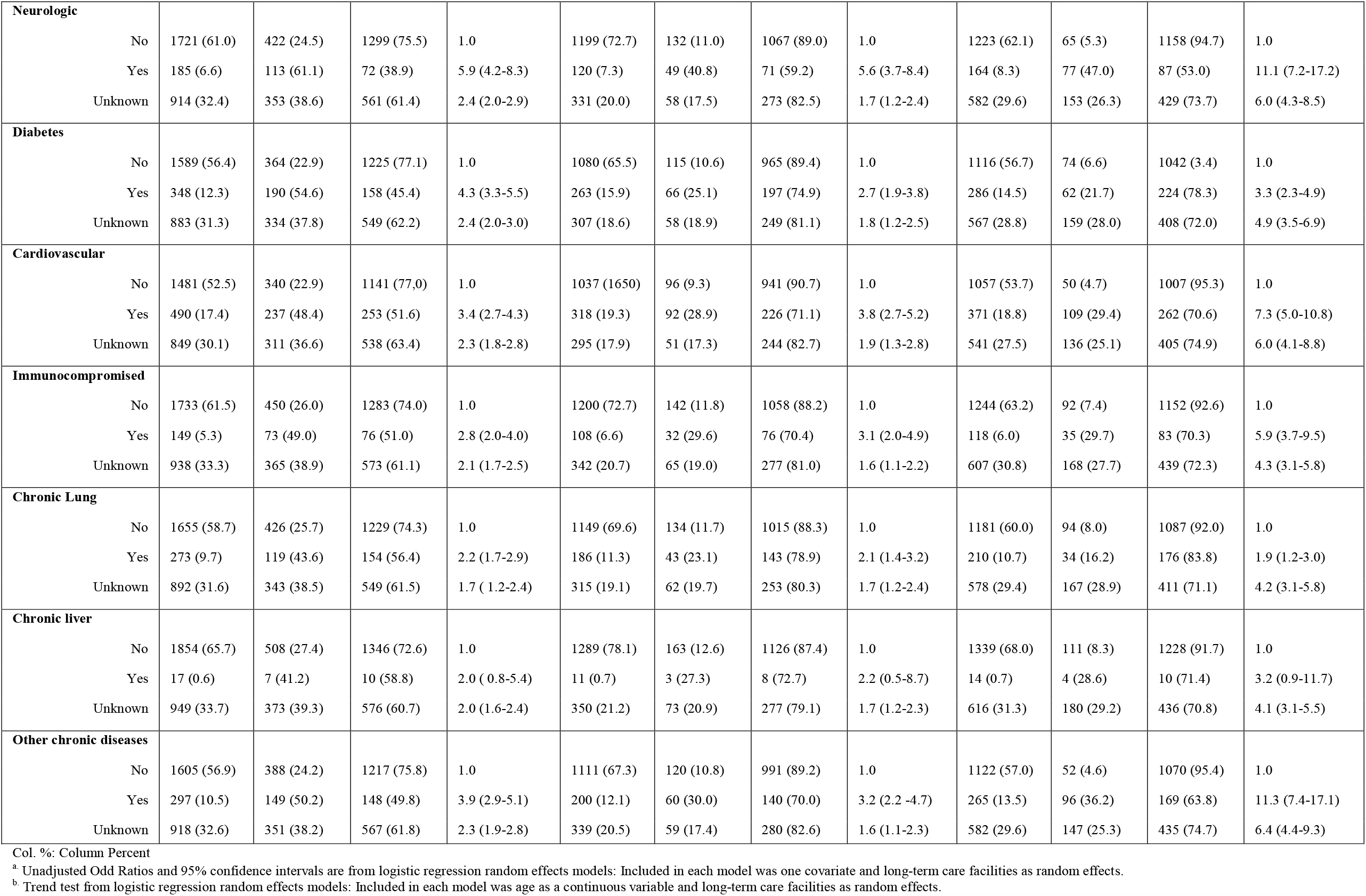
Characteristics of COVID-19 patients in Fulton County according to Hospitalization, ICU admission, and Death.

|  | Hospitalization-related information |  |  |  | ICU-related information |  |  |  | Mortality-related information |  |  |  |
| --- | --- | --- | --- | --- | --- | --- | --- | --- | --- | --- | --- | --- |
|  | Total | Hospitalized | Not Hospitalized |  | Total | Admitted to ICU | Not Admitted to ICU |  | Total | Died | Survived |  |
|  | No. (col. %) | No. (row %) | No. (row %) | OR (95% CI)<sup>a</sup> | No. (col. %) | No. (row %) | No. (row %) | OR (95% CI)<sup>a</sup> | No. (col. %) | No. (row %) | No. (row %) | OR (95% CI)<sup>a</sup> |
| **Total (%)** | 2820 (100.0) | 888 (31.5) | 1932 (68.5) |  | 1650 (100.0) | 239 (14.5) | 1411 (85.5) |  | 1969 (100.0) | 295 (15.0) | 1674 (85.0) |  |
| **Age** |  |  |  |  |  |  |  |  |  |  |  |  |
| Less than 25 | 204 (7.2) | 18 (8.8) | 186 (91.2) | 1.0 | 123 (7.5) | 4 (3.3) | 119 (96.7) | 1.0 | 130 (6.6) | 2 (1.5) | 128 (98.5) | 1.0 |
| 25-34 | 384 (13.6) | 62 (16.1) | 322 (83.9) | 2.0 (1.2-3.5) | 245 (14.8) | 13 (5.3) | 232 (94.7) | 1.7 (0.5-5.2) | 250 (12.7) | 3 (1.2) | 247 (98.8) | 0.8 (0.1-4.6) |
| 35-44 | 413 (14.7) | 79 (19.1) | 335 (81.1) | 2.5 (1.5-4.4) | 283 (17.2) | 14 (4.9) | 269 (95.1) | 1.5 (0.5-4.8) | 288 (14.6) | 2 (0.7) | 286 (99.3) | 0.4 (0.1-3.1) |
| 45-54 | 428 (15.2) | 115 (26.9) | 313 (73.1) | 4.1 (2.4-6.9) | 265 (16.1) | 33 (12.5) | 232 (87.5) | 4.2 (1.5-12.2) | 278 (14.1) | 16 (5.8) | 262 (94.2) | 3.9 (0.9-17.5) |
| 55-64 | 481 (17.1) | 172 (35.8) | 309 (64.2) | 6.9 (4.1-11.7) | 308 (18.7) | 44 (14.3) | 264 (85.7) | 5.0 (1.7-14.1) | 339 (17.2) | 33 (9.7) | 306 (90.3) | 6.5 (1.5-27.6) |
| 65-74 | 399 (14.1) | 180 (45.1) | 219 (54.9) | 13.0 (7.6-22.2) | 202 (12.2) | 57 (28.2) | 145 (71.8) | 11.7 (4.1-33.2) | 303 (15.4) | 69 (22.8) | 234 (77.2) | 19.2 (4.6-80.0) |
| 74 and above | 499 (17.7) | 260 (52.1) | 239 (47.9) | 22.6 (13.0-39.1) | 224 (13.6) | 74 (33.0) | 150 (67.0) | 14.6 (5.2-41.3) | 374 (19.0) | 170 (45.5) | 204 (54.5) | 54.4 (13.0-227.2) |
| Unknown | 12 (0.4) | 2 (16.7) | 10 (83.3) | 5.4 (1.0-28.0) | 0 (0.0) | 0 (0.0) | 0 (0.0) |  | 7 (0.4) | 0 (0.0) | 7 (100.0) |  |
| Median age (IQR) | 54 (37-69) | 64 (50-77) | 49 (34-63) | <0.0001<sup>b</sup> | 51 (36-65) | 66 (53-77) | 48 (35-62) | <0.0001<sup>b</sup> | 56 (38-70) | 77 (67-87) | 51 (36-65) | <0.0001<sup>b</sup> |
| **Gender** |  |  |  |  |  |  |  |  |  |  |  |  |
| Female | 1467 (52.0) | 415 (28.3) | 1052 (71.7) | 1.0 | 839 (50.8) | 89 (10.6) | 750 (89.4) | 1.0 | 997 (50.6) | 132 (13.2) | 865 (86.8) | 1.0 |
| Male | 1353 (48.0) | 473 (35.0) | 880 (65.0) | 1.4 (1.2-1.6) | 811 (49.2) | 150 (18.5) | 661 (81.5) | 2.1 (1.6-2.8) | 972 (49.4) | 163 (16.8) | 809 (83.2) | 1.8 (1.4-2.4) |
| **Race/ethnicity** |  |  |  |  |  |  |  |  |  |  |  |  |
| White (non-Hispanic) | 644 (22.8) | 152 (23.6) | 492 (76.4) | 1.0 | 389 (23.6) | 32 (8.2) | 357 (91.8) | 1.0 | 458 (23.2) | 63 (13.8) | 395 (86.2) | 1.0 |
| Black (non-Hispanic) | 1640 (58.2) | 632 (38.5) | 1008 (61.5) | 2.0 (1.6-2.5) | 969 (58.7) | 180 (18.6) | 789 (81.4) | 2.5 (1.6-3.7) | 1194 (60.6) | 219 (18.3) | 975 (81.7) | 1.4 (1.0-2.0) |
| Hispanic (all races) | 279 (9.9) | 68 (24.4) | 211 (75.6) | 1.0 (0.7-1.4) | 159 (9.6) | 17 (10.7) | 142 (89.3) | 1.5 (0.8-2.7) | 160 (8.1) | 9 (5.6) | 151 (94.4) | 0.6 (0.3-1.2) |
| Asian (non-Hispanic) | 61 (2.2) | 15 (24.6) | 46 (75.4) | 1.0 (0.6-1.9) | 41 (2.5) | 7 (17.1) | 34 (82.9) | 2.5 (1.0-6.1) | 38 (1.9) | 2 (5.3) | 36 (94.7) | 0.5 (0.1-2.3) |
| Other | 51 (1.8) | 9 (17.6) | 42 (82.4) | 0.7 (0.3-1.5) | 27 (1.6) | 2 (7.4) | 25 (92.6) | 1.0 (0.2-4.4) | 34 (1.7) | 1 (2.9) | 33 (97.1) | 0.3 (0.0-2.1) |
| Unknown | 145 (5.1) | 12 (8.3) | 133 (91.7) | 0.3 (0.2-0.5) | 65 (3.9) | 1 (1.5) | 64 (98.5) | 0.2 (0.0-1.4) | 85 (4.3) | 1 (1.2) | 84 (98.8) | 0.1 (0.0-0.6) |
| **Chronic renal** |  |  |  |  |  |  |  |  |  |  |  |  |
| No | 1752 (62.1) | 437 (24.9) | 1315 (75.1) | 1.0 | 1219 (73.9) | 136 (11.2) | 1083 (88.8) | 1.0 | 1244 (63.2) | 85 (6.8) | 1159 (93.2) | 1.0 |
| Yes | 157 (5.6) | 109 (69.4) | 48 (30.6) | 9.8 (6.5-14.8) | 103 (6.2) | 40 (38.8) | 63 (61.2) | 4.7 (3.0-7.3) | 139 (7.0) | 49 (35.3) | 90 (64.7) | 6.3 (4.0-10.0) |
| Unknown | 911 (32.3) | 342 (37.5) | 569 (62.5) | 2.2 (1.8-2.7) | 328 (19.9) | 63 (19.2) | 265 (80.8) | 1.8 (1.2-2.5) | 586 (29.8) | 161 (27.5) | 425 (72.5) | 4.6 (3.4-6.4) |

|  |  |  |  |  |  |  |  |  |  |  |  |  |  |
| --- | --- | --- | --- | --- | --- | --- | --- | --- | --- | --- | --- | --- | --- |
| **Neurologic** | No | 1721 (61.0) | 422 (24.5) | 1299 (75.5) | 1.0 | 1199 (72.7) | 132 (11.0) | 1067 (89.0) | 1.0 | 1223 (62.1) | 65 (5.3) | 1158 (94.7) | 1.0 |
|  | Yes | 185 (6.6) | 113 (61.1) | 72 (38.9) | 5.9 (4.2-8.3) | 120 (7.3) | 49 (40.8) | 71 (59.2) | 5.6 (3.7-8.4) | 164 (8.3) | 77 (47.0) | 87 (53.0) | 11.1 (7.2-17.2) |
|  | Unknown | 914 (32.4) | 353 (38.6) | 561 (61.4) | 2.4 (2.0-2.9) | 331 (20.0) | 58 (17.5) | 273 (82.5) | 1.7 (1.2-2.4) | 582 (29.6) | 153 (26.3) | 429 (73.7) | 6.0 (4.3-8.5) |
| **Diabetes** | No | 1589 (56.4) | 364 (22.9) | 1225 (77.1) | 1.0 | 1080 (65.5) | 115 (10.6) | 965 (89.4) | 1.0 | 1116 (56.7) | 74 (6.6) | 1042 (3.4) | 1.0 |
|  | Yes | 348 (12.3) | 190 (54.6) | 158 (45.4) | 4.3 (3.3-5.5) | 263 (15.9) | 66 (25.1) | 197 (74.9) | 2.7 (1.9-3.8) | 286 (14.5) | 62 (21.7) | 224 (78.3) | 3.3 (2.3-4.9) |
|  | Unknown | 883 (31.3) | 334 (37.8) | 549 (62.2) | 2.4 (2.0-3.0) | 307 (18.6) | 58 (18.9) | 249 (81.1) | 1.8 (1.2-2.5) | 567 (28.8) | 159 (28.0) | 408 (72.0) | 4.9 (3.5-6.9) |
| **Cardiovascular** | No | 1481 (52.5) | 340 (22.9) | 1141 (77.0) | 1.0 | 1037 (1650) | 96 (9.3) | 941 (90.7) | 1.0 | 1057 (53.7) | 50 (4.7) | 1007 (95.3) | 1.0 |
|  | Yes | 490 (17.4) | 237 (48.4) | 253 (51.6) | 3.4 (2.7-4.3) | 318 (19.3) | 92 (28.9) | 226 (71.1) | 3.8 (2.7-5.2) | 371 (18.8) | 109 (29.4) | 262 (70.6) | 7.3 (5.0-10.8) |
|  | Unknown | 849 (30.1) | 311 (36.6) | 538 (63.4) | 2.3 (1.8-2.8) | 295 (17.9) | 51 (17.3) | 244 (82.7) | 1.9 (1.3-2.8) | 541 (27.5) | 136 (25.1) | 405 (74.9) | 6.0 (4.1-8.8) |
| **Immunocompromised** | No | 1733 (61.5) | 450 (26.0) | 1283 (74.0) | 1.0 | 1200 (72.7) | 142 (11.8) | 1058 (88.2) | 1.0 | 1244 (63.2) | 92 (7.4) | 1152 (92.6) | 1.0 |
|  | Yes | 149 (5.3) | 73 (49.0) | 76 (51.0) | 2.8 (2.0-4.0) | 108 (6.6) | 32 (29.6) | 76 (70.4) | 3.1 (2.0-4.9) | 118 (6.0) | 35 (29.7) | 83 (70.3) | 5.9 (3.7-9.5) |
|  | Unknown | 938 (33.3) | 365 (38.9) | 573 (61.1) | 2.1 (1.7-2.5) | 342 (20.7) | 65 (19.0) | 277 (81.0) | 1.6 (1.1-2.2) | 607 (30.8) | 168 (27.7) | 439 (72.3) | 4.3 (3.1-5.8) |
| **Chronic Lung** | No | 1655 (58.7) | 426 (25.7) | 1229 (74.3) | 1.0 | 1149 (69.6) | 134 (11.7) | 1015 (88.3) | 1.0 | 1181 (60.0) | 94 (8.0) | 1087 (92.0) | 1.0 |
|  | Yes | 273 (9.7) | 119 (43.6) | 154 (56.4) | 2.2 (1.7-2.9) | 186 (11.3) | 43 (23.1) | 143 (78.9) | 2.1 (1.4-3.2) | 210 (10.7) | 34 (16.2) | 176 (83.8) | 1.9 (1.2-3.0) |
|  | Unknown | 892 (31.6) | 343 (38.5) | 549 (61.5) | 1.7 (1.2-2.4) | 315 (19.1) | 62 (19.7) | 253 (80.3) | 1.7 (1.2-2.4) | 578 (29.4) | 167 (28.9) | 411 (71.1) | 4.2 (3.1-5.8) |
| **Chronic liver** | No | 1854 (65.7) | 508 (27.4) | 1346 (72.6) | 1.0 | 1289 (78.1) | 163 (12.6) | 1126 (87.4) | 1.0 | 1339 (68.0) | 111 (8.3) | 1228 (91.7) | 1.0 |
|  | Yes | 17 (0.6) | 7 (41.2) | 10 (58.8) | 2.0 (0.8-5.4) | 11 (0.7) | 3 (27.3) | 8 (72.7) | 2.2 (0.5-8.7) | 14 (0.7) | 4 (28.6) | 10 (71.4) | 3.2 (0.9-11.7) |
|  | Unknown | 949 (33.7) | 373 (39.3) | 576 (60.7) | 2.0 (1.6-2.4) | 350 (21.2) | 73 (20.9) | 277 (79.1) | 1.7 (1.2-2.3) | 616 (31.3) | 180 (29.2) | 436 (70.8) | 4.1 (3.1-5.5) |
| **Other chronic diseases** | No | 1605 (56.9) | 388 (24.2) | 1217 (75.8) | 1.0 | 1111 (67.3) | 120 (10.8) | 991 (89.2) | 1.0 | 1122 (57.0) | 52 (4.6) | 1070 (95.4) | 1.0 |
|  | Yes | 297 (10.5) | 149 (50.2) | 148 (49.8) | 3.9 (2.9-5.1) | 200 (12.1) | 60 (30.0) | 140 (70.0) | 3.2 (2.2-4.7) | 265 (13.5) | 96 (36.2) | 169 (63.8) | 11.3 (7.4-17.1) |
|  | Unknown | 918 (32.6) | 351 (38.2) | 567 (61.8) | 2.3 (1.9-2.8) | 339 (20.5) | 59 (17.4) | 280 (82.6) | 1.6 (1.1-2.3) | 582 (29.6) | 147 (25.3) | 435 (74.7) | 6.4 (4.4-9.3) |

Col. %: Column Percent
<sup>a</sup>. Unadjusted Odds Ratios and 95% confidence intervals are from logistic regression random effects models: Included in each model was one covariate and long-term care facilities as random effects.
<sup>b</sup>. Trend test from logistic regression random effects models: Included in each model was age as a continuous variable and long-term care facilities as random effects.

**Figure 1.**
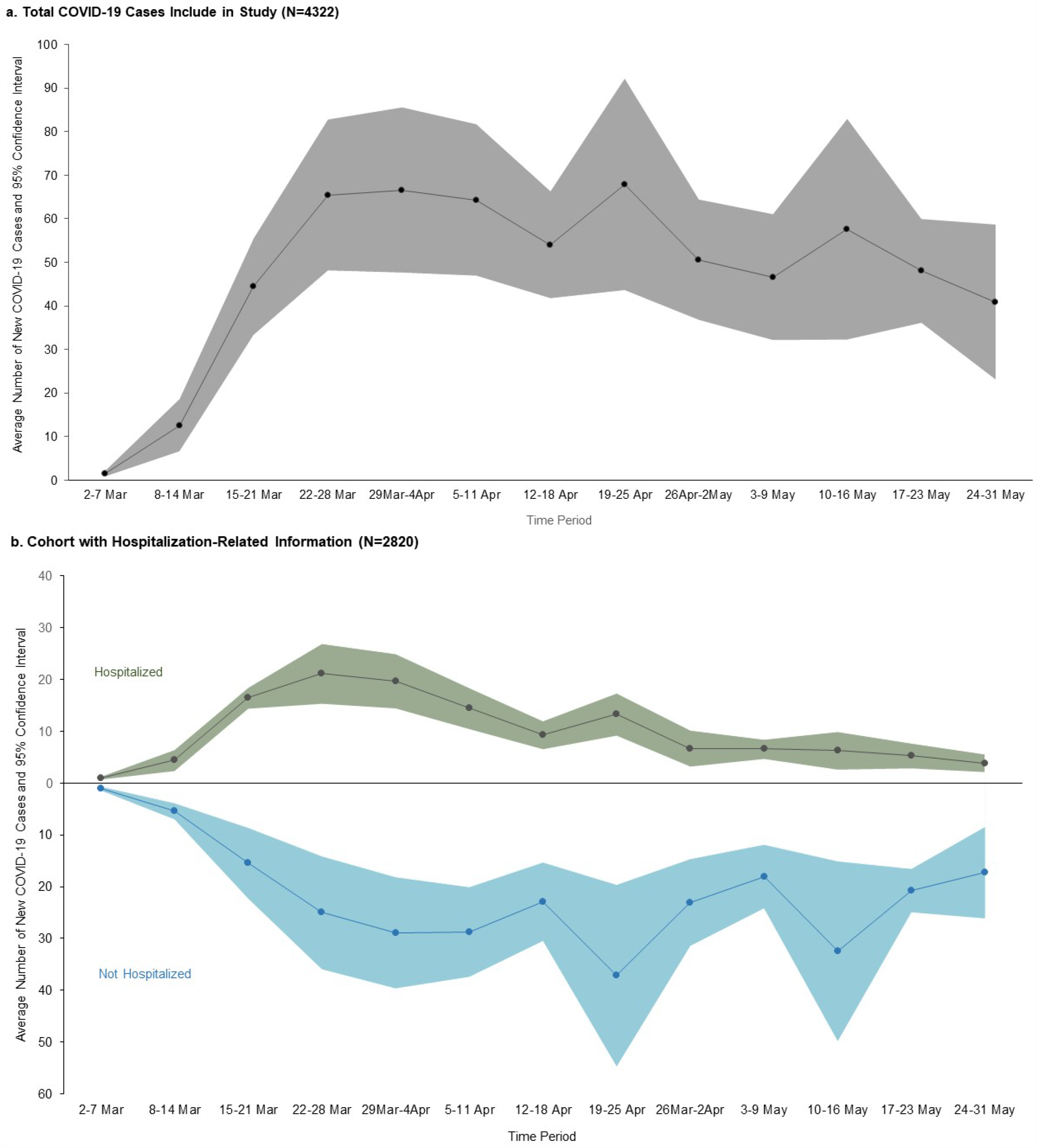
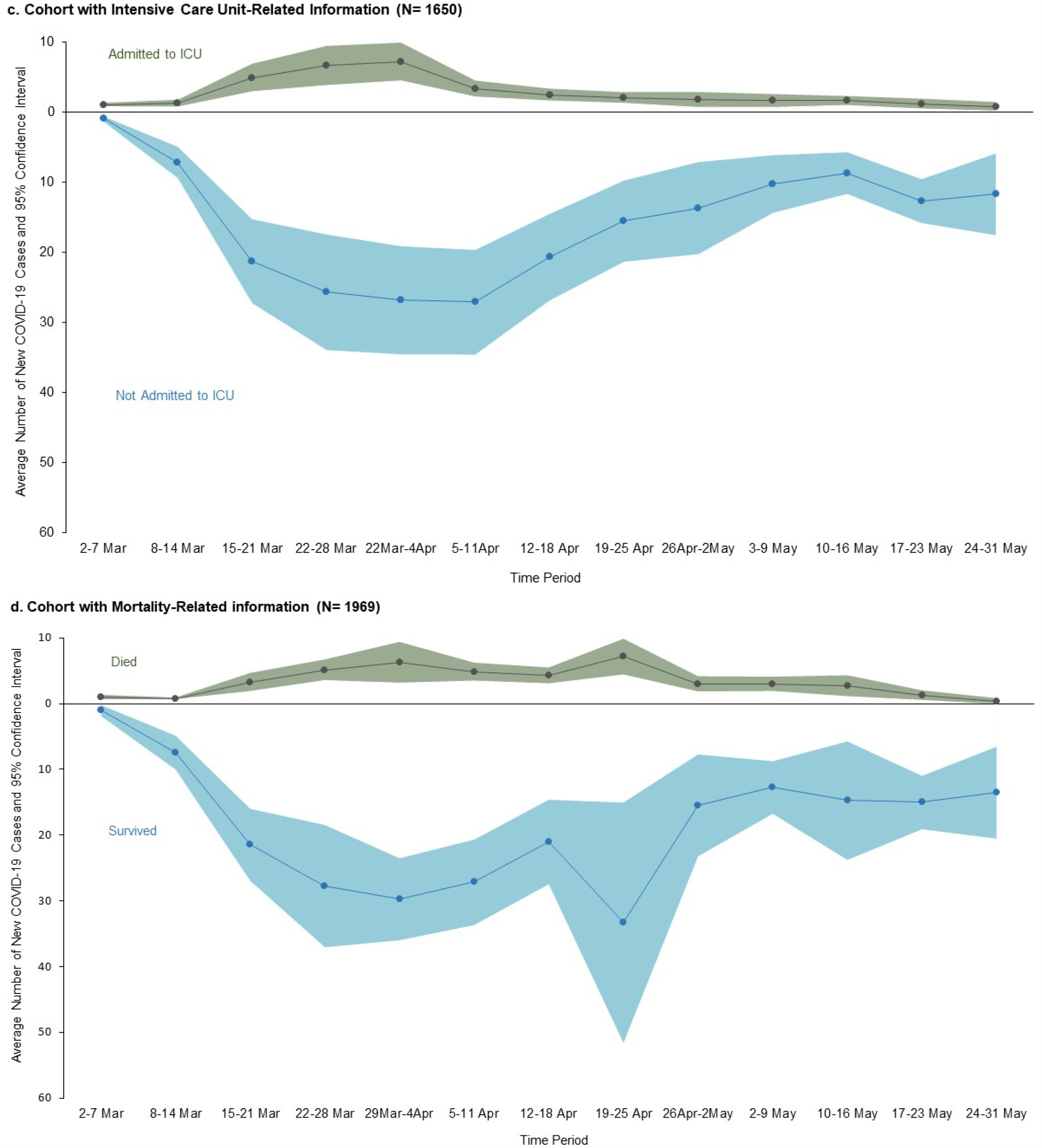
Average Number of New COVID-19 Cases and 95% Confidence Interval per Time Period in Fulton County (March 2–May 31, 2020) for the (a) Total Cohort Included, (b) Cohort with Hospitalization-Related Information, (c) Cohort with Intensive Care Unit-Related Information, and (d) Cohort with Mortality-Related Information.

### Risk Factor Analysis

In the unadjusted random effects models, age (≥25 years for hospitalization, ≥45 years for ICU admission, and ≥55 years for death), male gender and non-Hispanic black race were all significantly associated with being hospitalized, admitted to ICU. Similarly, age (≥25 years for hospitalization, ≥45 years for ICU admission, and ≥55 years for death) and male gender were associated with death, in exception for non-Hispanic black race which was not associated with death but was included in the multivariable models *a priori* (**Table 1**). Statistically significant associations with hospitalization, ICU admission, and death were found with the following comorbidities: chronic renal disease, neurologic disease, diabetes, cardiovascular disease, immunocompromised, chronic lung disease, and “other chronic diseases” (**Table 1**).

In the multivariable random effects models, we found that older patients were more likely to be hospitalized (age categories ≥25 years), admitted to ICU (age categories ≥55 years), die (age categories ≥65 years), or have severe COVID-19 (age categories ≥25 years) compared to those <25 years (**Figure 2**). Each 10-year increment in age was associated with a progressive increase in odds of hospitalization or severe disease. Even persons 45–54 years old had 3-fold higher odds of hospitalization or severe COVID-19 disease compared to persons under age 25 after adjusting for covariates.

**Figure 2.**
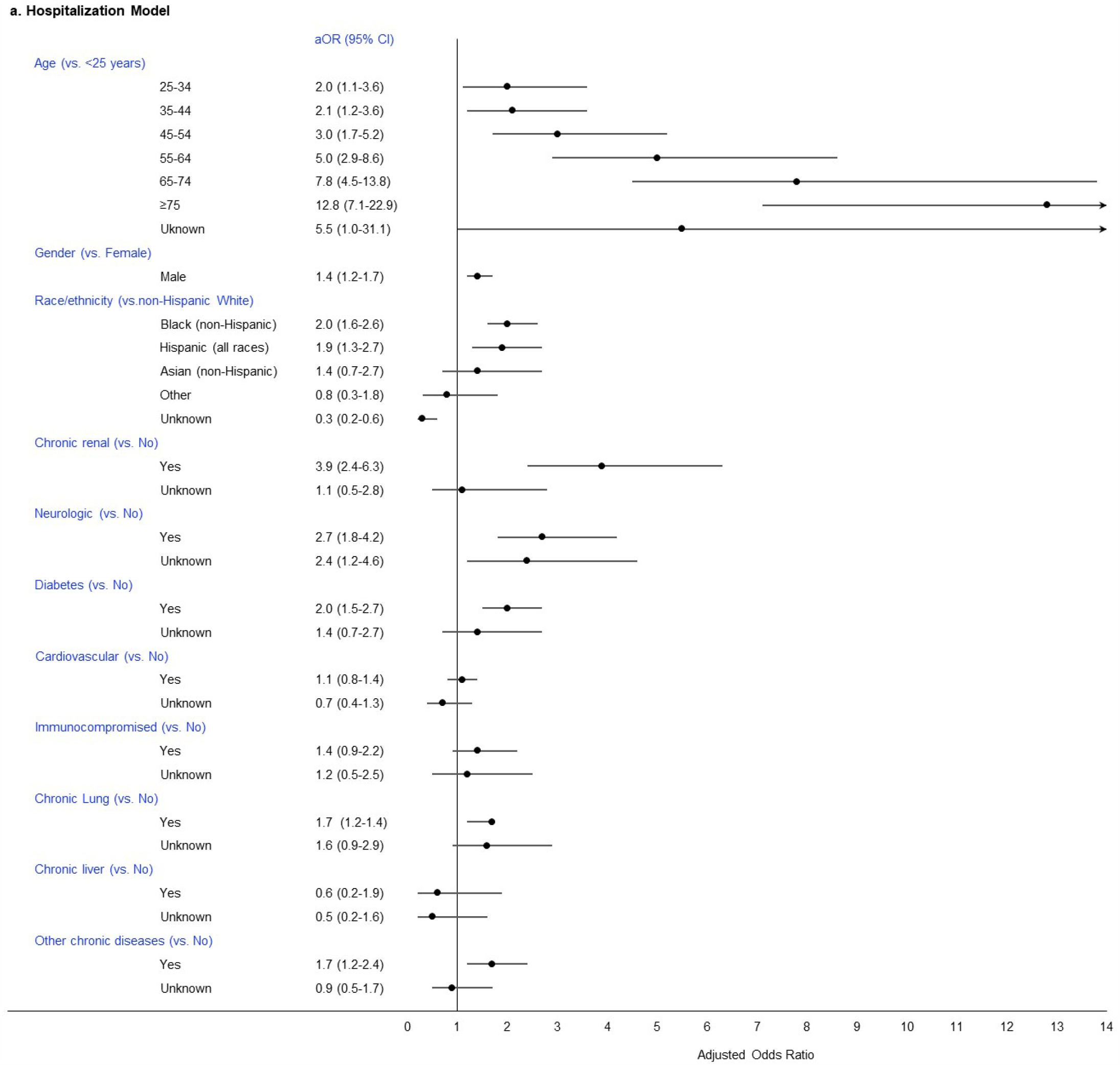
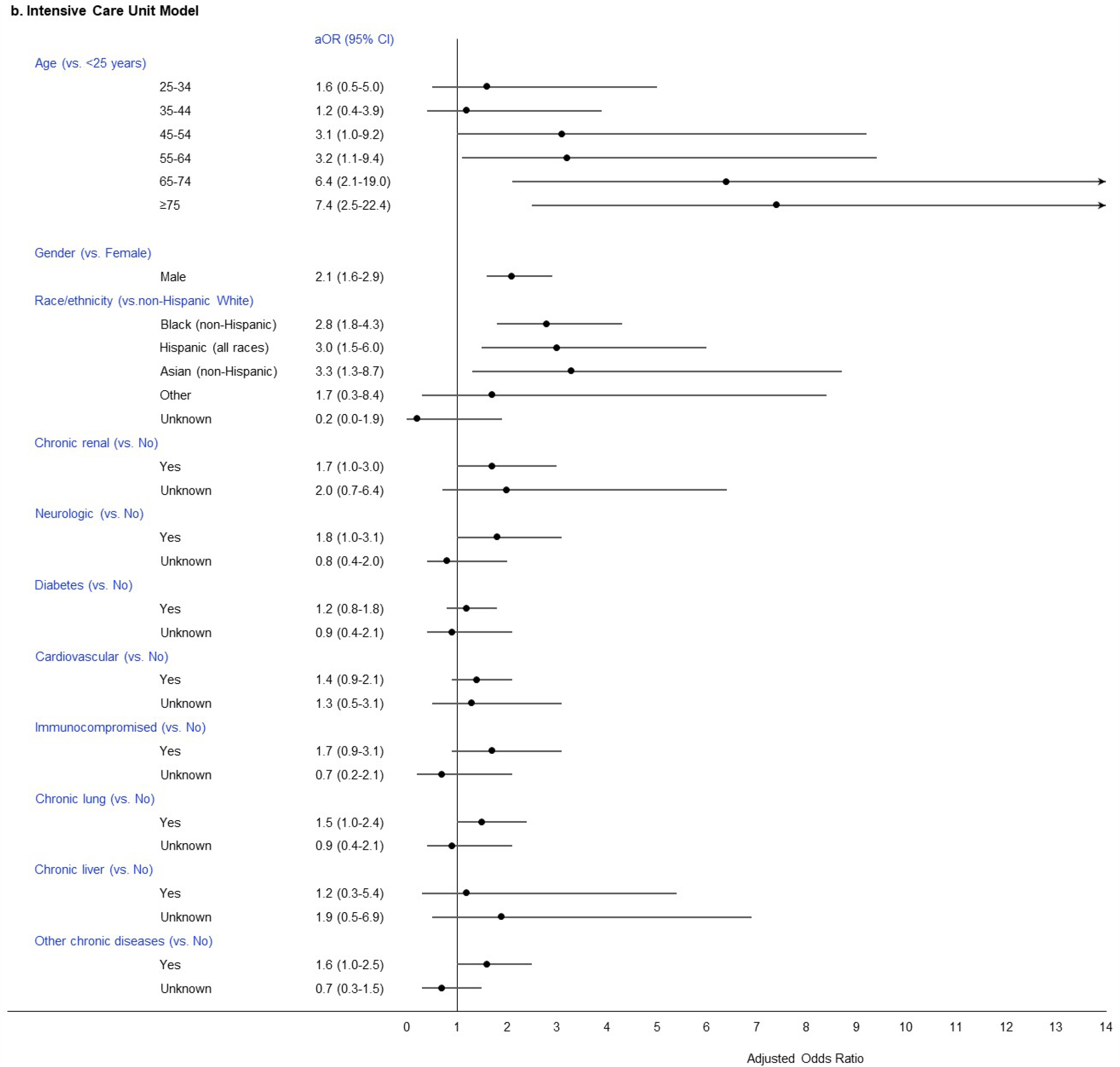
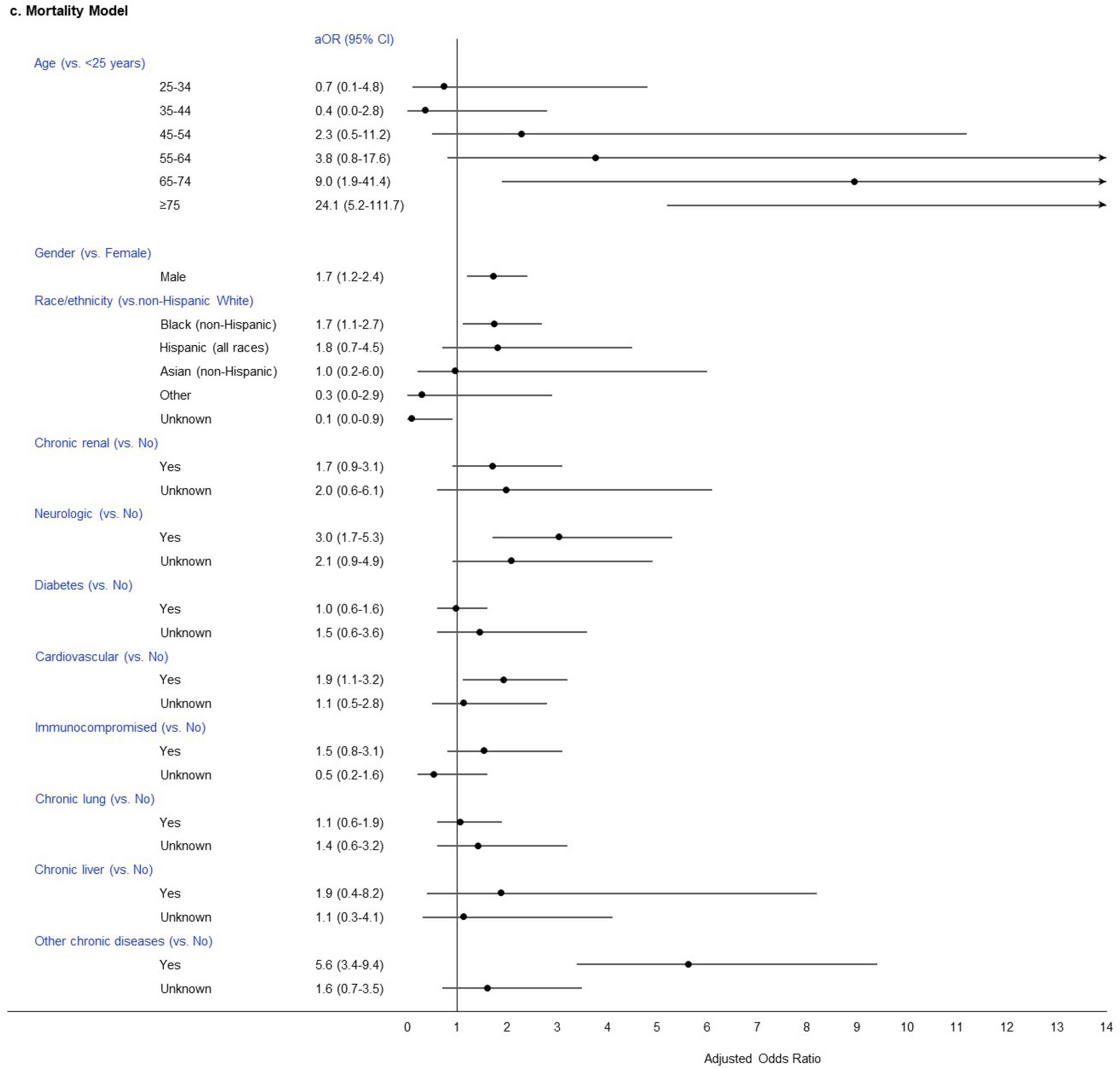
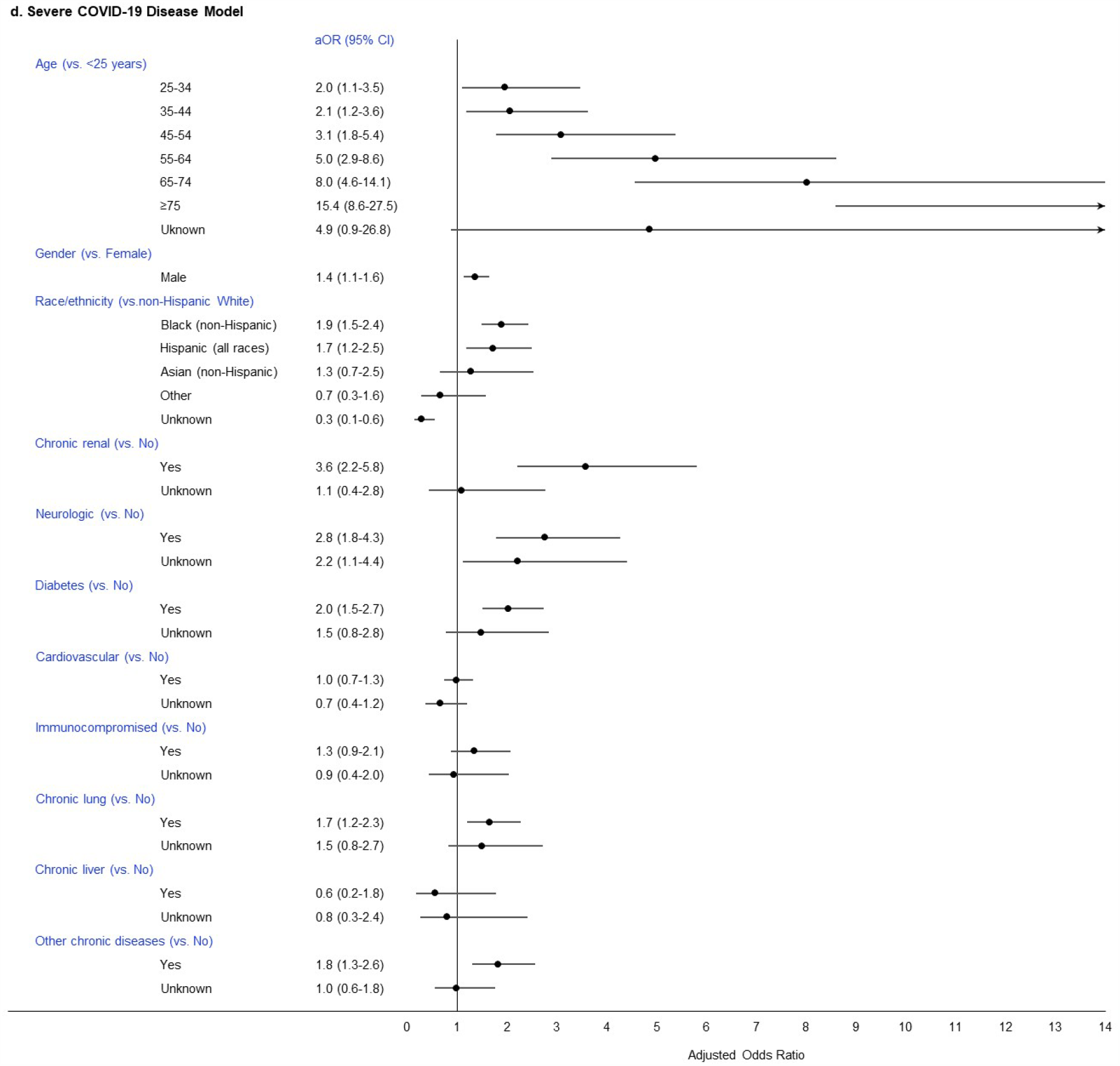
Multivariable Analyses of Risk Factors Associated with (a) Hospitalization, (b) ICU Admission, (c) Death, and (d) Severe Disease among Patients with Confirmed COVID-19 in Fulton County (March 2-May 31, 2020). aOR: adjusted odds ratio. The adjusted logistic regression random effects model included age categories, gender, chronic renal disease, neurologic disease, diabetes mellitus, cardiovascular disease, immunocompromised, chronic lung disease, chronic liver disease, and “other chronic diseases”, with long-term care facilities as random effects. Arrows pointing to the right for age indicate an upper 95% confidence limit that exceeds the odds ratio of 14.0 on the x-axis.

Males were more likely to be hospitalized (aOR 1.4, 95%CI: 1.2-1.7), admitted to the ICU (aOR 2.1, 95%CI: 1.6-2.9), die (aOR 1.7, 95%CI: 1.2-2.4) or have severe COVID-19 (aOR 1.4, 95%CI: 1.1-1.6) compared to females (**Figure 2**).

Patients who were non-Hispanic black (aOR 2.0, 95%CI: 1.6-2.6) and Hispanic (all races) (aOR 1.9, 95%CI: 1.3-2.7) were more likely to be hospitalized compared to patients who were non-Hispanic white (**Figure 2**). Similarly, non-Hispanic black and Hispanic cases had 2.8 and 3.0 higher odds of ICU admission, and 1.9 and 1.7 higher odds of severe COVID-19 disease, respectively, compared to non-Hispanic white cases. Non-Hispanic blacks also had 1.7 higher odds of death compared to non-Hispanic whites. This increase in the aOR for death to 1.7 for non-Hispanic blacks compared to the unadjusted OR of 1.4 indicates evidence of positive confounding by the adjusted covariates. Patients who were non-Hispanic Asian were more likely to be admitted to ICU (aOR 3.3, 95%CI: 1.3-8.7) compared to non-Hispanic whites, but their small numbers precluded estimates for other outcomes.

The composite outcome of “severe COVID-19” (**Figure 2d**) was more likely among patients with chronic renal disease (aOR 3.6, 95%CI: 2.2-5.8), neurologic disease (aOR 2.8, 95%CI 1.8-4.3), diabetes (aOR 2.0, 95%CI: 1.5-2.7), chronic lung disease (aOR 1.7, 95%CI: 1.2-2.3), or those with “other chronic disease” (aOR 1.8, 95%CI: 1.3-2.6) compared to those with no reported comorbidities.

We found evidence of interaction between race and gender in a random effects model that adjusted for age categories, gender, chronic renal disease, neural disease, diabetes mellitus, cardiovascular disease, immunocompromised state, chronic lung disease, chronic liver disease, and “other chronic diseases”. The odds of non-Hispanic black males having severe COVID-19 compared to non-Hispanic white males (aOR 3.5, 95%CI: 2.3-5.3) were higher than the odds of non-Hispanic black females having severe COVID-19 compared to non-Hispanic white females (aOR 1.6, 95%CI: 1.1-2.4) (likelihood ratio test for interaction, p <0.001).

In the sensitivity analysis, we found that each age group ≥25 years, male gender, non-Hispanic blacks, Hispanics (all races), having chronic renal disease, neurologic disease, diabetes, and having “other chronic diseases” all remained significantly associated with increased odds of having severe COVID-19 (**Figure 3**).

**Figure 3.**
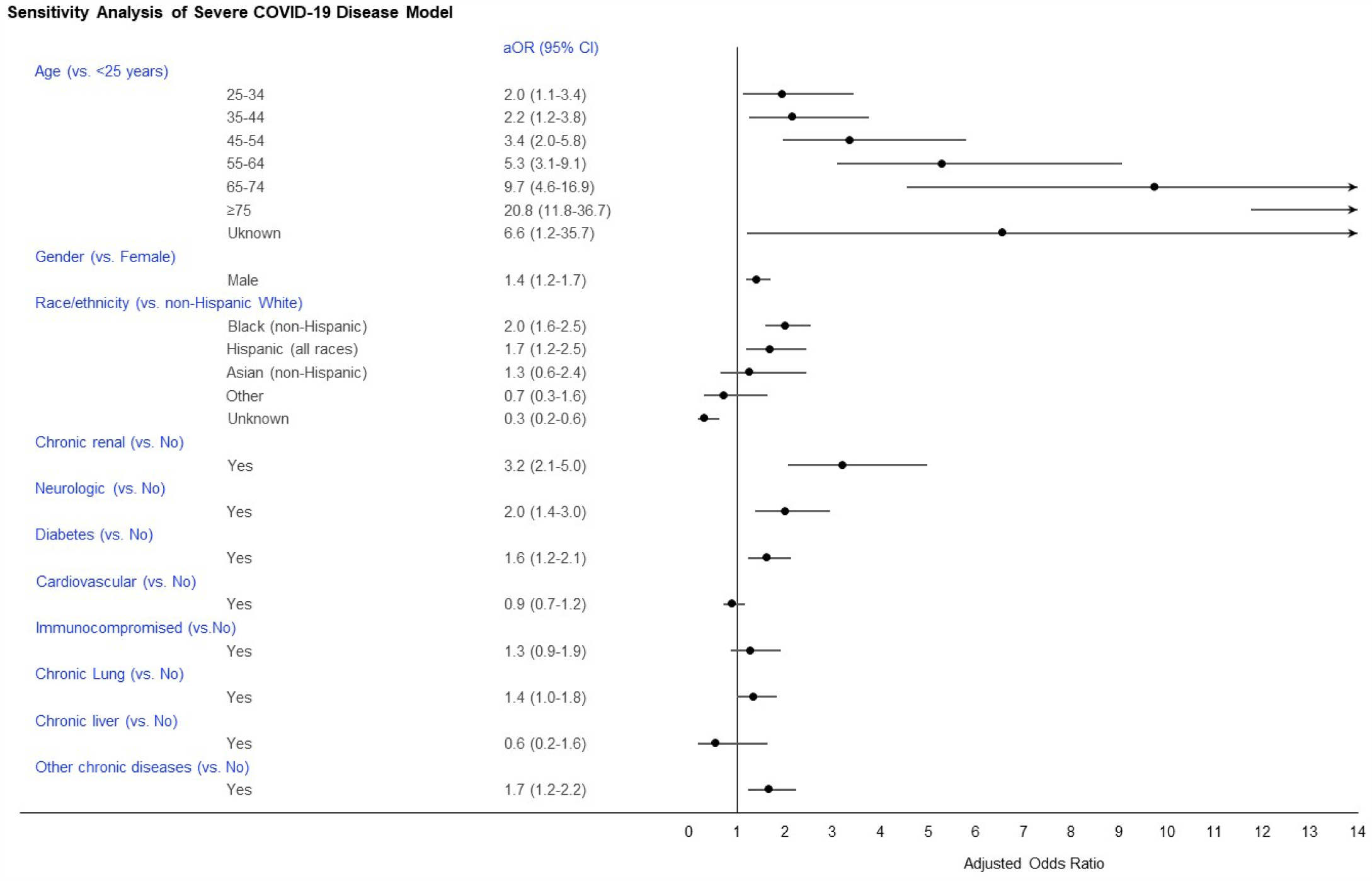
Sensitivity Analysis: Multivariable Analysis of Risk Factors Associated with Severe Disease among Patients with Confirmed COVID-19 in Fulton County (March 2-May 31, 2020). aOR: adjusted Odds Ratio. The adjusted logistic regression random effects model included age categories, gender, chronic renal disease, neural disease, diabetes mellitus, cardiovascular disease, immunocompromised state, chronic lung disease, chronic liver disease, and “other chronic diseases”, with long-term care facilities as random effects. Arrows pointing tp the right for age indicate a point estimate and/or upper 95% confidence limit that exceeds the odds ratio of 14.0 on the x-axis

## DISCUSSION

By July 2020, over 100,000 confirmed COVID-19 cases were reported in Georgia, suggesting that at a minimum, 1% of the population, and more likely 10% if we account for non or underdiagnoses, has been infected with SARS-CoV-2 in less than six months since the first case was reported.(14) In this study, we examined risk factors associated with hospitalization, ICU admission and death due to COVID-19 disease in Fulton County, Atlanta Metropolitan Area - the urban center of Georgia. We found that older age, male gender, black race and Hispanic ethnicity were consistently associated with these severe clinical outcomes, even after adjustment for comorbidities in multivariable analysis. Further, we found comorbid illnesses of diabetes, chronic renal disease, neurologic disease, chronic lung disease and other chronic disease were independently associated with the composite outcome of severe COVID-19 disease. These data provide valuable insights from the largest cohort of patients from the Southeast U.S., a region with rapidly rising rates of COVID-19, historically limited healthcare infrastructure, and widespread health disparities. Patients with these risk factors may benefit from proactive screening, higher prioritization for testing (if testing is limited), and earlier interventions to reduce morbidity and mortality from COVID-19.

At each step in the COVID-19 disease cascade (i.e., hospitalization; ICU; and death), we found a disproportionate burden among non-Hispanic blacks. Although they comprised 41% of all confirmed COVID-19 cases in Fulton County, 58% of cases that were hospitalized are black, and 73% of those who died are black. Importantly, the increased risk of hospitalization, ICU admission, or death among non-Hispanic black compared to non-Hispanic white patients is independent of age or co-morbid medical conditions. This difference was most pronounced among black men, who were at 3.5 times greater risk of having severe COVID-19 disease, though black women were still 1.6 times more likely than white women to have severe disease. We were unable to ascertain reasons for this. However, extensive research has demonstrated differences in health access and utilization as factors associated with poor outcomes for other infectious and chronic diseases in this population. Persons of Hispanic ethnicity similarly had a higher risk for severe disease; the smaller number of individuals in this group should not preclude public health efforts specifically targeted to diagnosis and prevention in these hard-hit communities. Indeed, persons of color may be at higher risk for SARS-CoV-2 exposure due to over-representation as workers in frontline industries.(15) Our findings add to literature from other parts of the U.S.(10, 16) that have highlighted racial disparities in COVID-19, but provides unique insights from an urban setting where COVID-19 outcomes have been better than what has been reported in other settings.(17)

Older age and comorbid medical conditions have been associated with more severe disease.(18-21) We found that risk of hospitalization and severe disease increased stepwise with every decade increase in age, with persons 25-34 years having 2-times the risk compared to those under age 25 years. Among persons 65 and older, the risk of severe disease was 8-15 times greater than among younger persons; this age group accounts for more than two-thirds of all deaths that occurred during the study period. However, recent shifts in testing have resulted in a rapid rise in case counts among persons who are 20-40 years old; by late-June, 2020 they comprised the majority of all diagnosed COVID-19 cases in Fulton County.(22) Thus, although they may be at only twice the risk as younger persons, this may translate into a large number of young adults experiencing severe disease. Consistent with trends across the U.S.,(23) we found a high prevalence of comorbid medical conditions among all COVID-19 cases, including 11.5% with cardiovascular disease, 8.2% with diabetes, and 7.0% with “other chronic disease” that were not individually recorded. Persons with diabetes, chronic renal disease, neurologic disease or chronic lung disease had at least double the risk of being hospitalized or having severe COVID-19 than those without these conditions, with chronic renal disease having the highest risk among all comorbidities. Importantly, in the stratified analysis by race/ethnicity, these comorbidities had an independent effect, not explained by higher prevalence among black or Hispanic individuals.

### Limitations

There were several limitations in our analysis. We utilized routinely collected public health surveillance data, which can have a reporting lag time of several weeks. To minimize this, we censored our data as of May 31, 2020, to allow sufficient time for complete reporting of key indicators, such as hospitalization and death. Data were incomplete for race, ethnicity, and comorbidities for persons whom we were unable to contact to complete a case investigation, which could be due to the absence of contact information or individuals declining to provide these data to the health department. These individuals may differ in important ways from those in whom we have complete information, including access to and utilization of healthcare services. However, our sensitivity analysis to account for missing data did not alter our results, supporting the robustness of the observed risk. Lastly, detailed information on clinical presentation and management of COVID-19 patients is not included in the surveillance database, limiting our ability to evaluate other factors that may be associated with severe disease.

## CONCLUSIONS

This study provides important insights into persons who at highest risk for severe COVID-19 disease in Fulton County – Atlanta Metropolitan Area. With expanded testing capacity and a greater understanding of COVID-19 pathophysiology and treatment options, these data can help guide public health programs and clinicians to better target testing and clinical interventions. As growing numbers of young people are being diagnosed with COVID-19, it is crucial to understand that the increased risk of severe disease starts as young as age 25 and that action in ensuring social distancing and return-to-work policy adequately protects this group. Lastly, the overwhelming data suggesting the highest risk of severe disease among non-Hispanic black men warrants particular attention and interventions designed to address this population that is simultaneously suffering from systemic inequities in overall health.

## Data Availability

Data were extracted from the State Electronic Notifiable Disease Surveillance System (SENDSS). SENDSS is an electronic database developed by the Georgia Department of Public Health that is used to track patients with notifiable diseases, including case investigation of COVID-19 patients across the state of Georgia.

https://sendss.state.ga.us/sendss/login.screen

## ARTICLE INFORMATION

### Author Contributions

All authors had full access to all the data in the study and takes responsibility for the integrity of the data and the accuracy of the data analysis.

*Concept and design:* Chishinga, Shah

*Acquisition, analysis, or interpretation of data:* All authors

*Drafting of the manuscript:* Chishinga, Shah

*Critical revision of the manuscript for important intellectual content:* All authors

*Statistical analysis:* Chishinga, Ghandhi, Shah

*Supervision:* Shah, Gandhi, F. Khan

### Conflict of Interest Disclosures

None reported.

### Funding/Support

Dr. Neel Gandhi was supported in part by funding from the US National Institutes of Health (K24AI114444, PI Gandhi).

### Disclaimer

The content of this report is solely the responsibility of the authors and does not represent the official position of Fulton County Board of Health, Fulton County Government, Brown University, or Emory University.

## Notes

### Competing Interest Statement

The authors have declared no competing interest.

